# Self-report inaccuracy in the UK Biobank: Impact on inference and interplay with selective participation

**DOI:** 10.1101/2023.10.06.23296652

**Authors:** Tabea Schoeler, Jean-Baptiste Pingault, Zoltán Kutalik

## Abstract

While the use of short self-report measures is common practice in biobank initiatives, such phenotyping strategy is inherently prone to reporting errors. In this work, we aimed to explore challenges related to self-report errors for biobank-scale research.

We derived a reporting error score (RE_SUM_) for n=73,129 UK Biobank (UKBB) participants, capturing inconsistent self-reporting in time-invariant phenotypes across multiple measurement occasions. We then performed genome-wide association scans on RE_SUM_, applied downstream analyses (LD Score Regression and Mendelian Randomization, MR), and compared its properties to a previously studied participation behaviour (UKBB participation propensity). The results were then used in extended analyses (simulations, inverse probability and variance weighting) to explore patterns and propose possible corrections for biases induced by reporting error and/or selective participation. Finally, to assess the impact of reporting error on SNP effects and trait heritability, we improved phenotype resolution for 15 self-report measures and inspected the changes in genomic findings.

Reporting error was present in the UKBB across all 33 assessed, time-invariant, measures, with repeatability levels as low as 11% (e.g., inconsistent recall of childhood sunburns). We found that reporting error was not independent from UKBB participation, evidenced by their negative genetic correlation (*r*_*g*_ = -0.90), their shared causes (e.g., education, income, intelligence; assessed in MR) and the loss in self-report accuracy following participation bias correction. Depending on where reporting error occurred in the analytical pipeline, its impact ranged from reduced power (e.g., for gene-discovery) to biased effect estimates (e.g., if present in the exposure variable) and attenuation of genome-wide quantities (e.g., 20% relative *h*^2^-attenuation for self-reported childhood height).

Our findings highlight that both self-report accuracy and selective participation are competing biases and sources of poor reproducibility for biobank-scale research. Implementation of approaches that aim to enhance phenotype resolution while ensuring sample representativeness are therefore essential when working with biobank data.

## Introduction

Genomic research is often confronted with large-scale datasets containing error in the phenotypic measures, as data collection is optimized towards the recruitment of large numbers of people. To reduce participant burden, save resources and maximize sample size, recruitment schemes often favour minimal phenotyping (i.e., the administration of short self-report scales) over precision phenotyping (i.e., the application of gold-standard measures). In the UK Biobank (UKBB), such self-report measures serve as the primary data source for commonly studied phenotypes, notably sociodemographic data, health-related information, behavioral outcomes, and lifestyles. While all phenotypes are measured with some degree of error, including those objectively ascertained (e.g., biological measures/laboratory assays), error induced by brief self-report measures pose a particular challenge when studying the associations of those phenotypes with genetic or other phenotypic information. As the reported information is influenced by subjective interpretation, misreporting, or cognitive limitations, error in self-report measures constitutes a potentially greater threat to the validity of findings.

While the early stages of genome-wide research were dominated by a push towards ever-growing sample sizes, challenges related to phenotype ascertainment are increasingly recognized as a non-negligible source of bias in genomic research^1,2^. While random error in phenotypes does not lead to bias in SNP estimates (cf., **sFigure 1**), the resulting measurement imprecision and increased Type-II error rates constitute one of the causes for large sample size requirements in genomic research. If gene-discovery is the primary study aim, increasing sample sizes can compensate for random error in the phenotype within the limits of feasibility. However, more problematically, random error puts an upper bound on how much variance can be explained in the phenotype^34^. Downstream genome-wide analyses focusing on variance components (e.g., heritability estimates^5^, polygenic prediction^6–8^) would therefore show (downward) bias in the presence of self-report inconsistencies.

Detecting and correcting self-report errors can be challenging when relying on biobank-scale data, as the required validation data is rarely available. However, with the increasing availability of repeated measurements in the UKBB, it is now possible to more systematically explore causes and consequences of self-report inconsistencies across measurement occasions. In this work, we aim to contribute to the growing body of research scrutinizing the impact of study design characteristics and participant behaviour (e.g., sampling procedures^9–11^, missing data^12^, study engagement^13^, data quality^14–16^) on findings obtained from biobank-scale data. Here, we focus on the challenges related to reporting error, defined as inconsistent self-reporting across measurement occasions. To that end, we aim to quantify error in commonly studied UKBB phenotypes, explore underlying characteristics and links with other participation behaviours, and assess its impact on genome-wide quantities. Such work is not only crucial for the interpretation of findings obtained from existing biobanks, but may help shape strategies aiming to enhance phenotype resolution in future biobank initiatives.

## Methods

### Indexes of reporting error in the UK Biobank

The UK Biobank is a large prospective study assessing more than 500,000 participants aged between 39 and 60 years who attended one of the baseline assessment centres between 2006 and 2010^17^. We first screened all UKBB phenotypes that could be used as indexes of reporting error, defined as inconsistent self-reporting over time. To that end, we included phenotypes that were assessed longitudinally but represented time-invariant variables, namely those that cannot change following the baseline assessment (e.g., self-reported birth weight, number of older siblings, age at first sexual intercourse). For each of the included time-invariant phenotypes, we partitioned its variance into its error-free and reporting error component, by regressing time point two phenotype (PT2, e.g., self-reported birth weight at follow-up) onto time point one phenotype (PT1, e.g., self-reported birth weight at baseline). Follow-up time (time between P_T1_ and P_T2_, time_T2-T1_) was included as a covariate in this model (P_T2_ = P_T1_ + time_T2-T1_). The variance explained by the model (*R*^*2*^) was used as an index of phenotype repeatability, such that 1-*R*^2^ quantifies the level of reporting error per phenotype. For comparison, we also estimated *R*^2^ for phenotypes subject to within-person temporal variability (including only objectively ascertained phenotypes, e.g., BMI, LDL) and measures subject to both temporal variability and reporting error (e.g., self-reported alcohol use, physical activity).

Next, to explore some of the properties underlying reporting error, we derived individual reporting error scores using a two-stage protocol; in stage one, we extracted the residuals (|RES_i_|) from a model regressing P_T2_ on P_T1_. In stage 2, the scaled residuals (|RES_i_|/ SD_T1,T2_) from stage one model were residualized for follow-up time (time_T2-T1_). The reporting error scores were then used as input for Principal Component Analysis (PCA) to obtain a weighted reporting error summary score. In PCA, we included only reporting error scores with at least 50,000 non-missing repeated observations. After combining the selected scores, we imputed missing values using row-wise mean imputation and performed PCA. Based on the first principal component, we then generated the weighted summary scores from the values of their observed indicator items. This score is a (weighted) average of reporting errors, representing the overall inaccuracy an individual exhibits when responding to time-invariant questions repeated over time. The resulting summary scores were used as the primary outcome in downstream analyses exploring correlates and causes of reporting error.

### Genome-wide analyses

The reporting error summary score (RE_SUM_) was then subjected to a genome-wide scan. For all genome-wide analyses (GWA), we restricted the sample to individuals of European ancestry based on principal components and excluded individuals with high missing rate and high heterozygosity on autosomes. Genetic variants were filtered according to Hardy-Weinberg disequilibrium (*P* > 1 × 10^-15^), minor allele frequency (> 1%), minor allele count (>100) and call rate (> 90%). The association tests were performed in REGENIE v2.0.2 (ref^18^), adjusting for age, sex and the first ten principal components. The resulting RESUM summary statistics file was then included in LD score regression^19^ (as implemented in GenomicSEM^20^) to estimate SNP heritability and genetic correlations with other traits. Genetic correlations were estimated for 39 publicly selected traits with available summary statistics files, where the selected traits tapped into participation behaviours (e.g., the UKBB participation probability, re-contact availability in the UKBB), physical features (e.g., height, body mass index), biological markers (e.g., LDL, systolic blood pressure), lifestyles (e.g., smoking, coffee intake), social variables (e.g., socioeconomic status, education), and mental health/personality (e.g., schizophrenia, ADHD, neuroticism) (cf. **sTable 1** in Supplement for details and references). To identify causal factors contributing to reporting error, we performed Mendelian Randomization (MR) as implemented in the R-Package TwoSampleMR21. Here, we used the same 39 selected traits with publicly available summary statistics files to extract genetic instruments for the exposure, where we selected LD-independent (--clump-kb 10,000 --clump-r2 0.001) SNPs reaching genome-wide significance (*p*<5×10^-8^). We only performed MR for exposures with at least five genetic instruments. Tests of causality were performed using the inverse-variance weighted (IVW) MR estimator, where the reporting error GWA output was included as the outcome. To facilitate comparability of the results, we standardized the SNP effects (βSTD) prior to conducting MR. βSTD per SNP *j* was obtained by dividing the *z*-score per SNP [Z_j_=β(SNP_j_)/SE(SNP_j_)] by the square root of the sample size [βSTD(SNP_j_)=Z_j_/√N]. The results were corrected for multiple testing using FDR correction (controlled at 5%), correcting for the total number of performed tests per downstream analysis (LDSC and MR).

### Assessing the link between reporting error and UK Biobank participation

To explore patterns of covariation between reporting error other participatory behaviours that are known to bias genome-wide estimates, we also included ‘UKBB participation probabilities’ in the analytical pipeline described above. This trait was derived as part of a previous study^10^ focusing on the impact of participation bias on genome-wide findings. In brief, the participation probabilities are the predicted probabilities of UKBB participation (with 1= individuals taking part in the UKBB and 0=individuals taking part in a representative reference sample), based on 14 harmonized demographic, social and lifestyle variables.

Phenotypically, we estimated the level of covariation between the reporting error summary score and the UKBB participation probability. In addition, we obtained the standardized coefficients of the 14 baseline variables predicting UKBB participation (representative sample = 0; UKBB= 1) as done in our previous work^10^, to compare the coefficients to those obtained when including the reporting error summary score as the outcome. The total variance explained by the 14 predictors was obtained from LASSO regression (fivefold cross-validation) in glmnet^22^, which also included all possible two-way interaction terms among the categorical (dummy) and continuous variables. To assess if UKBB participation and reporting error share similar genetic and causal structures, we applied the same genome-wide pipeline as described above (i.e., performing LDSC regression and MR analyses) to UKBB participation (n=283,749) as the outcome of interest. The summary statistic file from the GWA on UKBB participation is accessible via the GWAS catalogue (accession number GCST90267294).

Finally, within a regression framework, adjustment for selective participation (unequal inclusion probabilities) and reporting error (unequal error variances, heteroskedasticity) can be achieved by the implementation of weights, where over-represented/reporting error-prone individuals are down-weighted and under-represented/reporting error-free individuals are up-weighted. To assess how weighting informed by participation and/or reporting error affect phenotype and sample characteristics, we derived reporting error weights (*w*_*RE*_), indexed as the inverse of the error variance 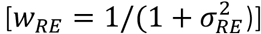. 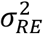 was obtained by taking the average of the reporting error variances (*Var*_*P*_) across the time-invariant phenotypes (P) selected for PCA: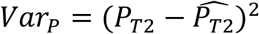, where 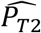 are the fitted values from a model regressing the standardized phenotype assessed at follow-up (*P*_*T*2_) on the standardized phenotype assessed at baseline (*P*_*T*1_). We then assessed changes in sample and phenotype characteristics following inverse probability/variance weighting, where we included either the UKBB participation weights (*w*_*P*_), the error weights (*w*_*RE*_) or the error-adjusted participation weights (*w*_*P×RE*_ =*w*_*P*_ *× w*_*P*_). Change was assessed at the level of (a) measurement repeatability in time-invariant phenotypes (i.e., comparing estimates of R^2^ obtained in an unweighted versus weighted sample) and (b) means in continuous phenotypes known to link to UKBB participation (i.e., comparing the weighted and unweighted means obtained for years of education and age).

### Simulations

To illustrate the individual and combined impact of reporting error and participation bias on exposure-outcome associations in a realistic setting, we simulated data for two phenotypes included in exposure-outcome linear regression models (education, BMI), the two participation behaviours of interest (reporting error, study participation), and modelled the relationships among these variables. The two phenotypes of interest, BMI and education, were chosen as these represent two continuous traits with different measurement properties (reporting error-free versus reporting-error prone measure, respectively) and have been linked to UK Biobank participation^10^.

The following simulation scenarios were tested: a) the ground truth, where the causal effect of the exposure on the outcome was estimated in a representative sample, and the exposure and outcome were measured without error, b) reporting error only scenario, where reporting error was present in the exposure or outcome measure (but no participation bias) c) participation bias only scenario, where we introduced participation bias (but no measurement error) and d) reporting error and participation bias scenario, where both reporting error and participation bias were introduced. These scenarios were then simulated within a bi-directional framework, testing the effects of (error-free) BMI on (error-prone) education and vice-versa. The data-generating mechanisms are depicted in the directed acyclic graphs (DAG) shown in **Figure 5**.

The coefficients used in the simulation scenarios were derived as follows from the UKBB data:

For UKBB participation, we used the standardized coefficients for education (*β*_*EDU*_) and BMI (*β*_*BMI*_) on UKBB participation as estimated in MR (described above). To obtain the coefficients required to simulate reporting error in self-reported years of education, we regressed the reporting error score for education (RES_EDU_, as described above) onto education (E) and BMI (B) and extracted the standardized effect estimates: *RES*_*EDU*_ = *α*_*EDU*_ *E* + *α*_*BMI*_ *B* + *∈*.

The obtained coefficients were then used to simulate the data, where biases were introduced as follows: for participation bias, we first generated the simulated participation probabilities,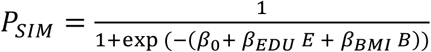, where *E* and *B* denote the simulated variables for years of education (E) and BMI (B), respectively. The variables were simulated as *E*∼ *N*(0,1) and *B* ∼ *N* (0,1) when included as the exposure and as *E* =*νB* +*∈* and *B* =*νE* +*∈* when included as the outcome, where *∈*∼*N*(0,1 – *ν*^2^) and *ν* denotes the true causal effect of the exposure on the outcome. The coefficient *β*_0_ was set to mimic the UKBB response rate, where around 5.5% of the 9,000,000 individuals initially invited to take part were recruited in the study^17^ 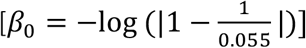. Subjects were then assigned a random number *U* from the uniform distribution *U* ∼ *Uniform*(0,1) and were classified as either respondent (U < P_SIM_) or nonrespondent (U > P_SIM_).

Reporting error was generated for one self-report measure (education, *E*), and was simulated as heteroskedastic error. Heteroskedasticity in this context refers to error in the measured phenotype (E_measured_) that is nonconstant and varies across individuals: *E*_*measured*_ =*E*_*true*_+∈_*EDU*_, where ∈_*EDU*_∼ N(0,*R*). *R* was simulated as *R*_*SIM*_ = *α*_*EDU*_ *E* +*α*_*BMI*_, *B* +*∈*, which was then scaled to have a standard deviation of 1 and values of *R* > 0[*R* =(*R*_*SIM*_ +|min(*R*_*SIM*_|)/*sd(R_SIM_*)]. BMI was modelled as an error-free measure in all simulation scenarios [*B*_*measured*_ = *B*_*true*_].

The impact of reporting error and selective participation was assessed in terms of bias (i.e., beta coefficients of the exposure-outcome association) and root-mean-square error (RMSE), an index that captures both the severity of the bias and the variance of the estimator: 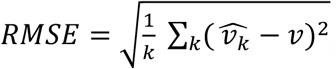, where 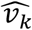 is the estimated effect of the exposure-outcome association at simulation kand *ν* is true causal effect of the exposure on the outcome. We performed k=1000 simulations and true causal effect was set to *ν* =−0.2.

### Impact of reporting error on SNP effects and trait heritability

To explore the impact of reporting error on genome-wide quantities, we compared the results from GWA tests on error-corrected versus error-prone versions of the same phenotype. We derived error-corrected phenotypes by taking the mean across multiple measurement occasions (e.g., mean in self-reported childhood height), as the within-person average reduces the random error in a variable. The baseline phenotype assessed in the same subset of UKBB participants was used as the error-prone counterpart (e.g., baseline self-reported childhood height). Genome-wide tests using REGENIE were then performed on both the repeated-measure and the single-measure phenotype. LD-independent SNPs reaching genome-wide significance (*p* < 5×10^−8^) were selected via clumping (clump-kb, 250; clump-r2, 0.1), and the explained variance per SNP *j* was obtained by squaring standardized beta (βSTD). We estimated SNP heritability for both the single-measure (*h*^*2*^S) and the repeated-measure GWA (*h*^*2*^R) and calculated the difference (*h*^*2*^*DIFF*= *h*^*2*^R - *h*^*2*^S) using the following test statistic:

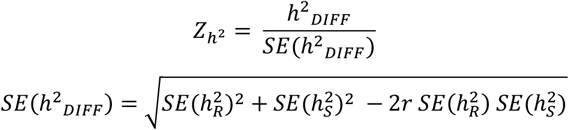

The correlation coefficient r(*h*^*2*^R, *h*^*2*^S) was obtained from 200-block jackknife analysis, where we split the genome into 200 equal blocks of SNPs and removed one block at a time to perform jackknife estimation. *h*^*2*^*DIFF* was obtained for traits with at least 2% SNP heritability.

## Results

### Indexes of reporting error in the UK Biobank

As shown in **Figure 1 (sTable 2**, Supplement), reporting error (RE) was present across all of the 33 assessed UK Biobank time-invariant phenotypes, with a mean error estimate of 0.232 [possible range: 0 (absence of error) to 1]. High levels of measurement repeatability were present for self-reports providing information about major life events, such as date of birth (R^2^>0.99), number of children (R^2^=0.99), country of birth (R^2^=0.99). A substantial proportion of self-reports showed questionable levels of repeatability, notably variables relying heavily on recall of childhood histories, such as childhood sunburns (R^2^=0.11) or comparative childhood body size (R^2^=0.47). **Figure 1** also illustrates the level of repeatability for variables containing error due to misreporting and/or temporal variability. Here, self-report measures subject to temporal instability showed particularly low levels of repeatability, notably diet (e.g., sodium and vitamin D intake in last 24 hours) and other lifestyles (e.g., physical activity in last 24 hours). Five UKBB phenotypes had data from directly comparable objective and subjective measures. Estimation of R^2^ revealed that the concordance between the two data sources (objective versus subjective) was low, ranging from R^2^=0.002 (vitamin D, self-report versus blood measure) over R^2^=0.031 (sleep, self-reported versus accelerometer derived) to R^2^=0.252 (first child’s birthweight, self-reported versus hospital records) (**sFigure 2** and **sTable 3**, Supplement).

**Figure 1.**
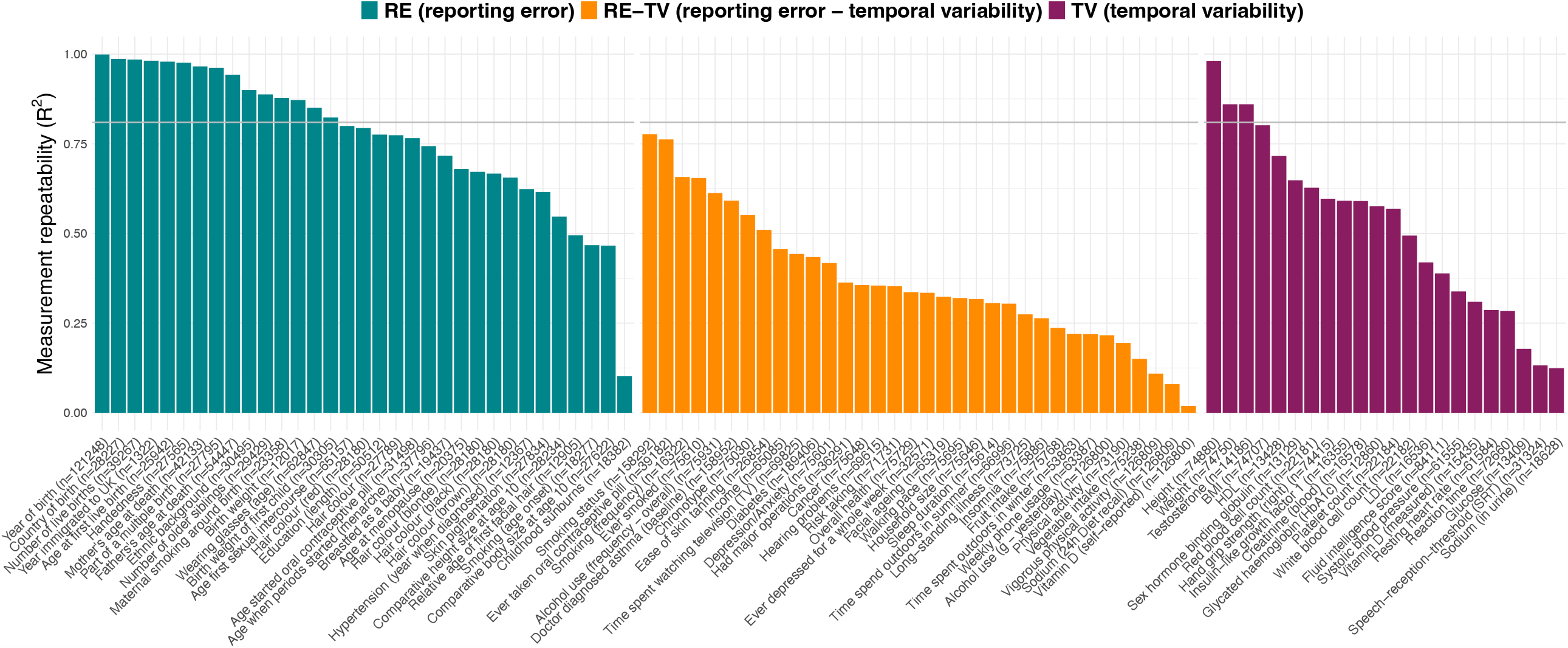
Measurement repeatability of UK Biobank self-report and objective measures R^2^= Variance explained by models regressing phenotype (P) measured at time point 2 (P_T2_, e.g., birth weight reported at follow-up) onto the phenotype assessed at time point 1 (P_T1_, e.g., selfreported birth weight assessed at baseline), while controlling for follow-up time (time_T2-T1_). Variables with R^2^ estimates above the grey line indicate variables with high levels of repeatability (R^2^ >0.9^2^).

Next, we generated the reporting error scores (RESi, illustrated in **Figure 2A**), indexing the level of reporting inconsistency per phenotype and UKBB participant. **sFigure3-4** (Supplement) summarize the contribution of baseline age, follow-up time, their interaction (age x follow-up time) and sex on the reporting error scores, highlighting that the scores varied mostly as a function of follow-up time and its interaction with age. In addition, reporting error was more prevalent among males, as 12 (70.59%) of the 17 reporting error scores showing significant sex-differential effects were higher in males than in females. The largest sex-differential effect was present for self-reported mother’s age at death, where females showed substantially lower levels of reporting error.

**Figure 2.**
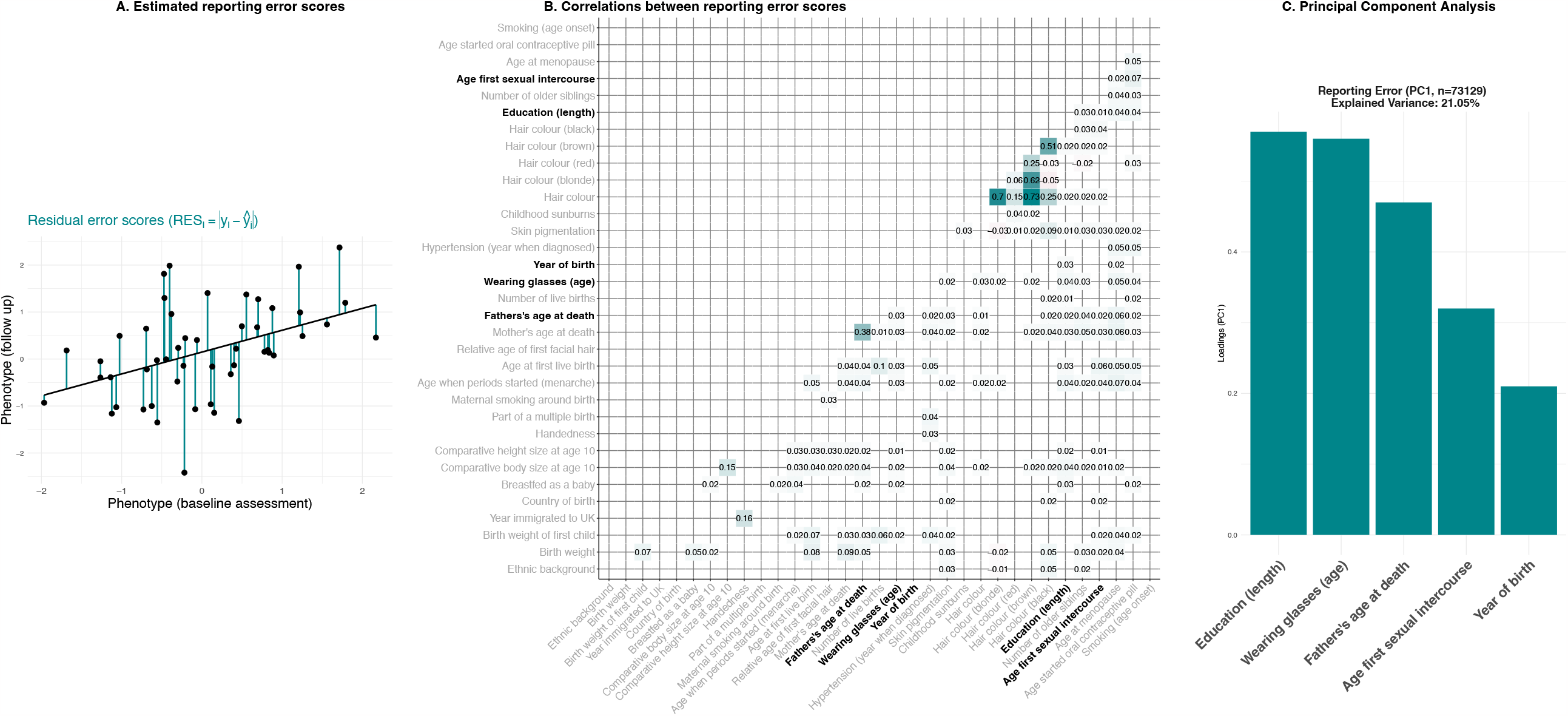
Weighted reporting error summary scores. Panel A. Illustration of a reporting error score for a particular phenotype, derived as the residual scores from a model regressing the phenotype measured at time point 2 (P_T2_, e.g., birth weight reported at follow-up) onto the phenotype assessed at time point 1 (P_T1_, e.g., self-reported birth weight assessed at baseline), controlled for follow-up time (time_T2-T1_). The reporting (residual) error scores are shown as the vertical deviations of the observed values (y_i_) around the fitted line. Panel B. Correlation matrix highlighting significant (*p*<0.05) Pearson correlation coefficients between the reporting error scores. Labels in bold highlight variables that were included in Principal Component Analysis. Panel C. Summary of results from Principal Component Analysis, highlighting the variance explained by the first principal component (PC1) and the loadings of the indicators on PC1.

Assessing the correlations among reporting error scores (**Figure 2B**), we found that the majority of correlations were small but positive [159 (96.36%) out of the 165 significant correlations]. The largest positive correlations were present among measures tapping into similar constructs, such as the r(mother’s age at death, father’s age at death)=0.37 or r(comparative body size at age 10, comparative height size at age 10)=0.15. Including five of the reporting error scores with n>50,000 in principal component analysis (years of education, age when started wearing glasses, father’s age at death, age at first sexual intercourse, year of birth), the first principal component (*PC*_1_) explained 21% of the variance. The individual reporting error scores all loaded positively on *PC*_1_ (**Figure 2C**).

### Assessing the link between reporting error and UK Biobank participation

To examine if reporting error varied as a function of sample representativeness, we first assessed the level of covariation between reporting error and UKBB participation. Phenotypically, we found a negative correlation (r_PEARSON_ = -0.10) between the reporting error summary score and UKBB participation, indicating that a greater willingness to participate in the UKBB links to more consistent self-reporting. Similarly, we observed negative genetic correlations between reporting error and other participatory behaviours, including the UKBB participation probability (r_g_ = -0.86, 95%CI -1.01; -0.72), re-contact availability in the UKBB (r_g_ = -0.73, 95%CI -0.88; -0.58) and follow-up (mental health survey) participation (r_g_ = -0.64, 95%CI -0.79; -0.49) (cf. **Figure 4** and **sTable 4**).

To assess shared and non-shared characteristics between reporting error and UKBB participation, we then tested for associations between a number baseline characteristics and the two outcomes (**Figure 3A, sTable 5**). Here, significant predictors differentially linked to the two outcomes, where female participants with higher levels of education and lower BMI showed less reporting errors but a higher willingness to take part in the UKBB. Only age predicted the two outcomes in the same direction, such that older individuals tended to show more reporting errors and were also more likely to participate in the UKBB. Including all predictors simultaneously in LASSO regression explained around 12% of the variance in UKBB participation and 6% in reporting error.

**Figure 3.**
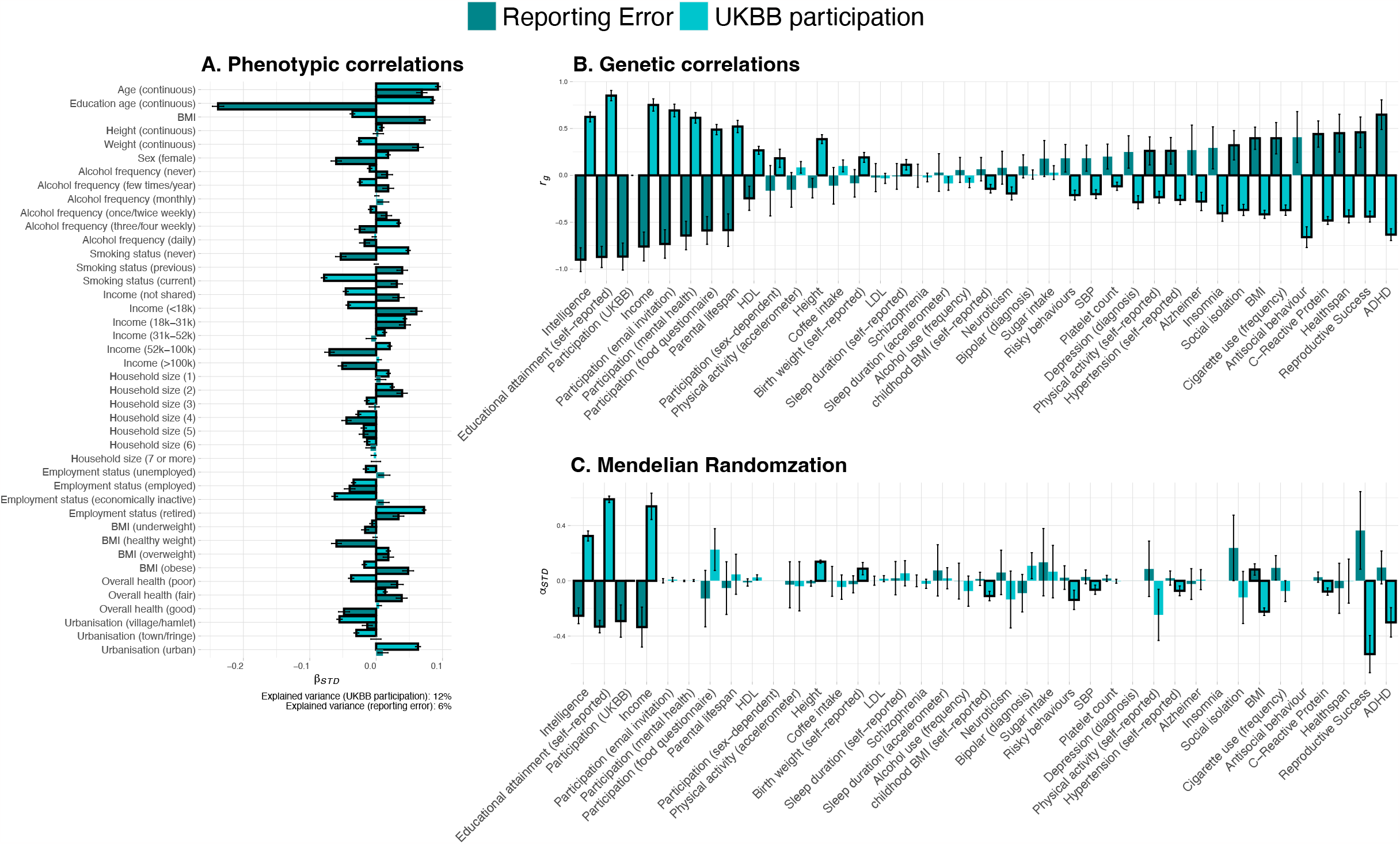
Correlates and causes of reporting error and UKBB participation **Panel A.** Standardized coefficients (and 95% confidence intervals) of variables predicting reporting error (in dark turquoise) and UKBB participation (in light turquoise) in univariate regression models. **Panel B.** Genetic correlations (r_g_) and corresponding 95% confidence intervals of reporting error (n =62,131) and UKBB participation (n = 283,749) with other traits. Significant genetic correlations (*p*_FDR_<0.05) are highlighted with black borders. **Panel C.** Standardized estimates (α_std_) obtained from Mendelian Randomization analyses on reporting error and UKBB participation as the outcomes. Significant MR estimates (*p*_FDR_<0.05) are highlighted with black borders.

The weighted reporting error summary scores (RE_SUM_) showed low but significant levels of SNP heritability (h^2^RESUM=2.63%, 95% CI 1.22%-4.04%). In line with the phenotypic correlations, reporting error and UKBB participation differentially correlated with most of the socio-educational and behavioural variables included in LD score regression (**Figure 3B, sTable 4**). These included intelligence (*rg*_Reporting_ = -0.9, *rg*_Participation_ =0.62), years of education (*rg*_Reporting_ = -0.87, *rg*_Participation_ = 0.85) and income (*rg*_Reporting_ = -0.76, *rg* _Participation_ = 0.75). Similarly, applying Mendelian Randomization analysis to identify causal factors contributing to reporting error, we find that reporting error and UKBB participation were explained by mostly socio-educational variables, where higher income, years of education and intelligence reduce self-report errors (standardized effect α_Income_ = -0.36, α_Education_ =-0.33, α_Intelligence_ = -0.25) but increase the probability of UKBB participation (α_Income_ =0.54, α_Education_ = 0.59, α_Intelligence_ =0.32) (**sTable 6**, Supplement).

**Figure 4** shows the distribution of the participation (inverse probability) weights and reporting error (inverse variance) weights. The performance of the inverse variance weights was assessed in terms of reporting error reduction in eight phenotypes, including those used in PCA and three additional phenotypes showing the largest degree of reporting error (**Figure 1**, i.e., body size at age 10, age when started smoking, number of childhood sunburns). Both the inverse variance weights and the participation weights performed as intended, in that they reduced the error variance in the eight variables inspected for measurement inconsistencies (i.e., increasing the level of measurement repeatability R^2^, **Figure 4B**) and made the sample more representative (i.e., lowering the mean age and mean level of education, **Figure 4C**), respectively. As the variability among the participation weights was large (indicating likely risk of bias due to selective participation), its application resulted in a substantial loss in effective sample size (62%, from n=63898 to n_EFF_ = 24,438, **Figure 4A**). In contrast, the reporting error weights showed little variability, causing a minimal loss in effective sample size (2%, from n=63,898 to 62,627). The reporting-error adjusted participation weights (inverse variance weights × participation weights) no longer reduced reporting error in all instances, and re-introduced a slight shift towards non-representativeness, resulting in a slight increase in effective sample size when compared to the unadjusted participation weights (24,438 versus 24,623).

**Figure 4.**
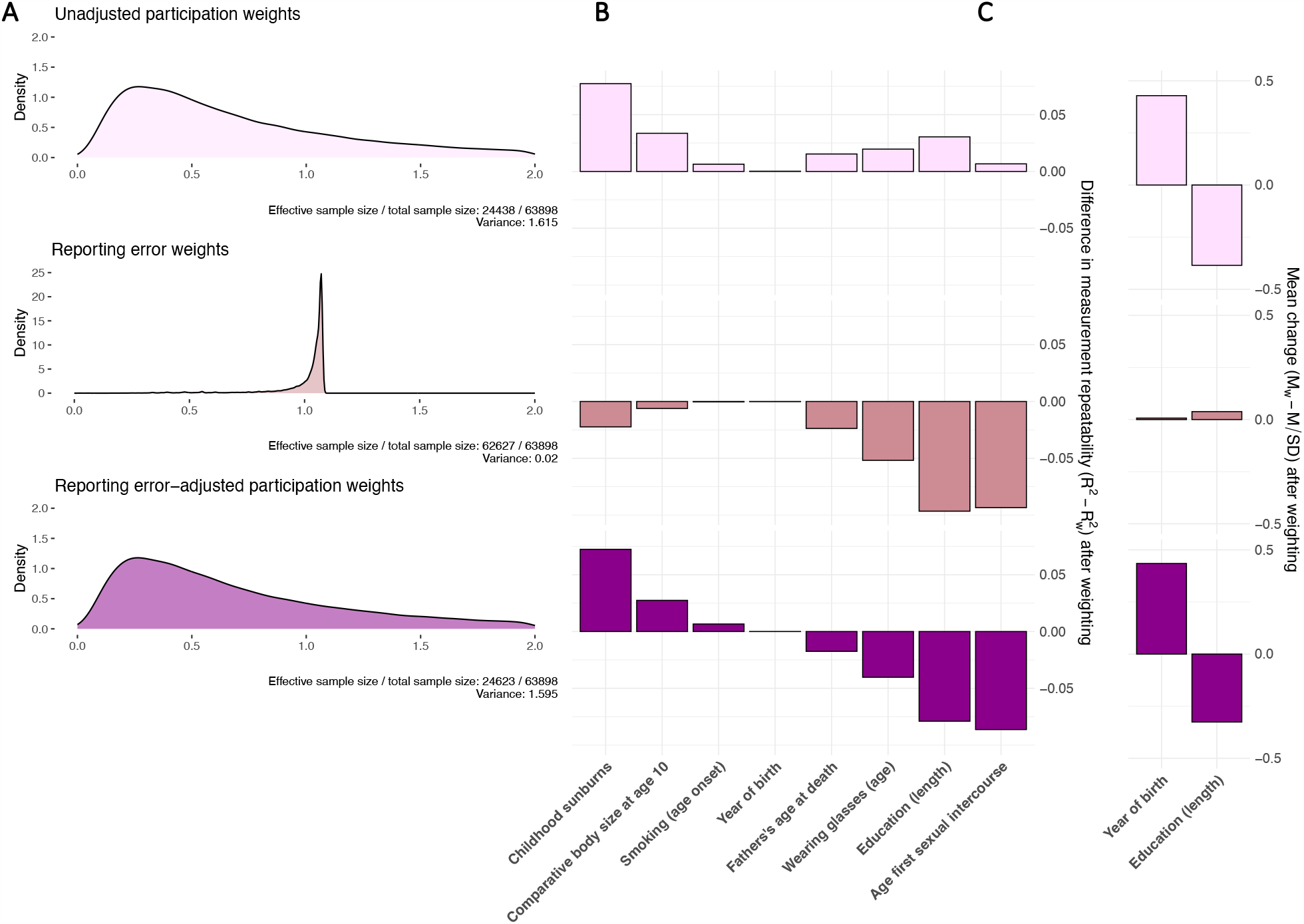
Reporting error-adjusted participation weights **Panel A.** Truncated density curves of the normalized UK Biobank weights (w), estimated for n=63,898 participants. The effective sample size was estimated as *n* ×{1/[Var(w) + 1]}. **Panel B.** R^2^=Variance explained by standard (ordinary least squares) regression models regressing phenotype (P) measured at time point 2 onto the phenotype assessed at time point 1, while controlling for follow-up time (timeT2-T1). R^2^_W_ = Variance explained by weighted (weighted least squares regression) models, incorporating UK Biobank weights to adjust for selective participation (top panel: unadjusted participation weights), reporting error (middle panel: reporting error weights) or both (bottom panel: reporting error-adjusted participation weights). Positive values in R^2^_DIFF_ (R^2^-R^2^_W_) index reduced measurement repeatability following weighting. **Panel C.** Change in means as a function of weighting, obtained for two continuous phenotypes known to link to UK Biobank participation (age, education). Change in means was expressed as a standardized mean difference, i.e., difference between the unweighted mean (*m*) and the weighted mean (*m*_w_), divided by the unweighted standard deviation (*m*_w_ − *m*/sd).

### Simulations

We tested eight simulation scenarios to illustrate the individual and combined impact of reporting error and selective participation on exposure-outcome associations (**Figure 5**). The following standardized beta coefficients for education and BMI on reporting error (R) and the participation probabilities (P) were estimated and used to simulate the data:

**Figure 5.**
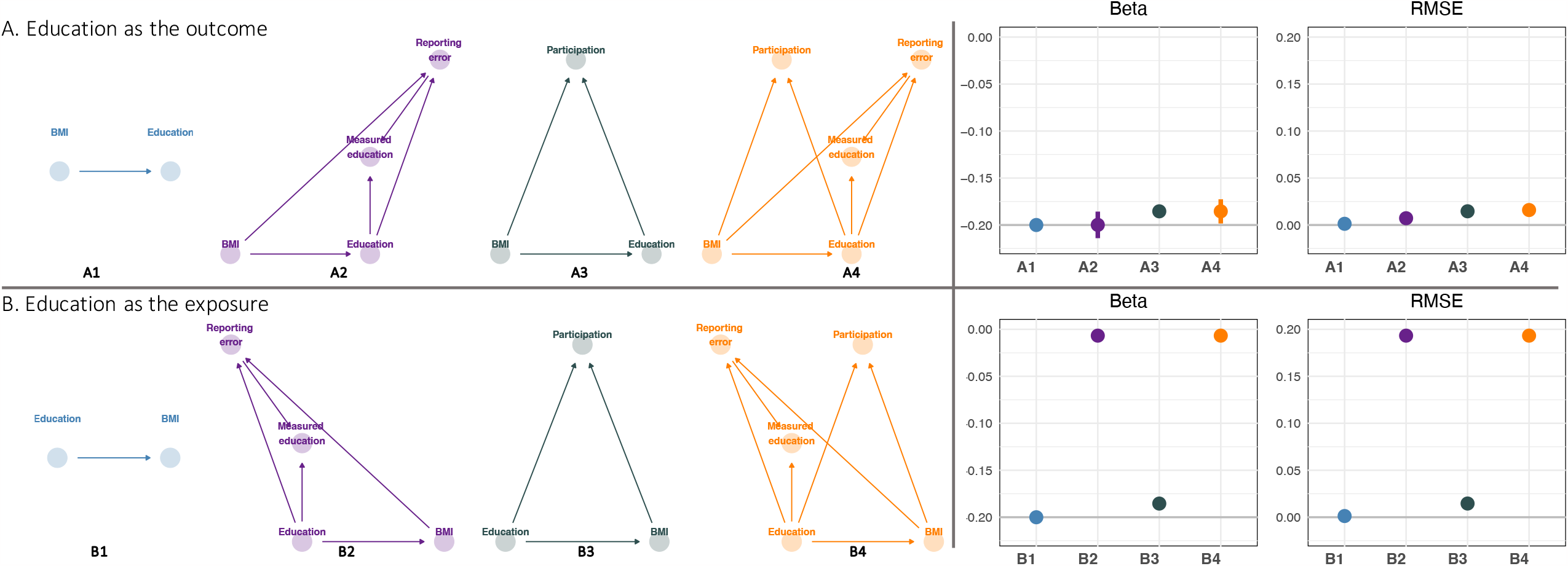
Simulations illustrating the impact of reporting error and/or selective participation on exposure-outcome associations Directed Acyclic Graphs (DAGs) illustrating the different simulation settings, where reporting error (DAGs highlighted in violet), participation bias (DAGs highlighted in green) or both (DAGs highlighted in orange) were present when assessing the effect of BMI on self-reported education (**panel A**) and the effect of self-reported education on BMI (**panel B**). The impact of the two participatory behaviours (reporting error, participation) was assessed in terms of in terms of bias (**panel C**, the beta coefficient of the exposure-outcome association) and root-mean-square error (RMSE, **panel D**). The true causal estimates was set to be -0.2 (grey line, **panel C**).

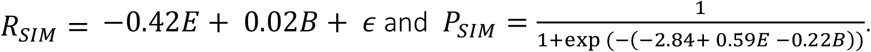

We found that deviations from the true causal effect resulted from both selective participation and reporting error in the exposure, in both cases leading to downward bias in the effect estimate (Panel C, **Figure 5**). RMSE was most strongly increased by reporting error in the exposure (Panel D, **Figure 5**), reflecting a large bias in the effect estimate towards the null. While reporting error in the outcome did not induce bias in the effect estimate, the increased uncertainty in parameter estimates also raised the RMSE, a measure that combines both bias and variance.

### Impact of reporting error on SNP effects and trait heritability

To assess the impact of reporting error on genome-wide results, we compared the output obtained from genome-wide analyses on single-measure phenotypes (e.g., self-reported childhood height assessed at baseline) versus repeated-measure phenotypes (using the average across multiple measurement occasions) (**Figure 6**). In total, 417LD-independent SNPs reached significance (*p* < 5×10^−8^) in genome-wide scans on the 12 traits, of which 79 (18.94%) were only identified in repeated-measure GWA. Among the identified SNPs, the explained variance increased following error-correction for 285 SNPs (68.35%). While the beta estimates obtained from the two sets of GWA were the same (**sTable 7**, Supplement), in accordance with the simulations demonstrating that reporting error in the outcome does not induce bias, the reduced error in the phenotype value narrowed the standard errors of the effect estimates, thereby boosting power for genome-wide discovery.

**Figure 6.**
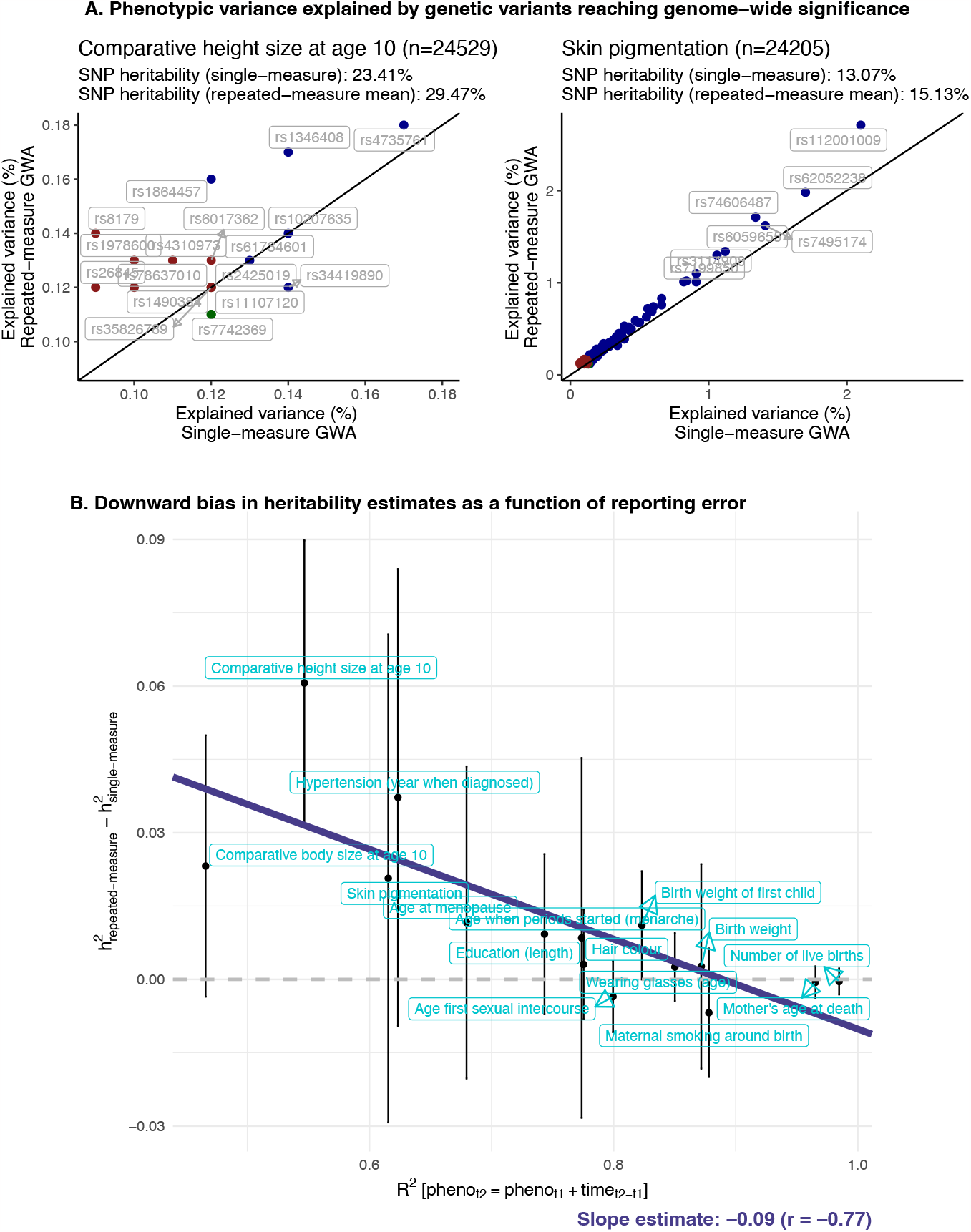
Impact of reporting error on SNP effects and trait heritability **Panel A.** Explained variance (β_STD_^2^) per SNP reaching genome-wide significance in error-corrected GWA analyses (y-axis, phenotype obtained using means across multiple measurement occasions) or error-uncorrected GWA analyses (x-axis, phenotype obtained from a single baseline measure). The colour scheme highlights in which GWA the genetic variant was identified, including error-corrected GWA (in red), error-uncorrected GWA (in green) or in both (blue). **Panel B**. The y-axis shows the differences in SNP heritability estimates obtained from error-corrected GWA analyses and error-uncorrected GWA analyses (h^2^_DIFF_ = h^2^_repeated-measure_ – h^2^_single-measure_). The x-axis gives the degree of repeatability per phenotype, estimates as the variance (R2) explained by models regressing phenotype (P) measured at time point 2 on the phenotype assessed at time point 1, while controlling for follow-up time (time_T2-T1_) and age.

Finally, with respect to SNP-based heritability estimates, we find that enhanced phenotype resolution increased *h*^*2*^ estimates. Overall, the degree of *h*^*2*^-disattenuation was proportional to the degree of reporting error per phenotype [r(h^2^DIFF, R^2^Repeatabiliy)= -0.77], where the largest notable downward bias in *h*^*2*^ estimates was present for self-reported height size at age 10 (R^2^Repeatabiliy=0.55, *h*^*2*^single-measure =23% versus *h*^*2*^repeated-measure=30%). The complete set of results is included in the Supplement (**sFigure 5, sTable 7-8**).

## Discussion

Phenotyping based on short self-report measures is common practice in biobank schemes, which has paved the way for large-scale genome-wide discovery studies involving millions of individuals. While such assessments are cost-effective and minimize the invested time of the participants, they are particularly prone to errors resulting from misreporting. In this study, we quantified the extend of reporting error for commonly studied UK Biobank (UKBB) phenotypes, assessed its properties and links with other participation behaviours, and evaluated its impact on exposure-outcome and genotype-phenotype associations. Overall, we found that reporting error is non-negligible for many commonly studied self-report measures, notably those relating to early life histories (e.g., puberty, education, childhood height/weight), common environmental exposures (e.g., number of sunburns) or lifestyles (e.g., age when started smoking). Consequently, exploiting large biobank samples does not necessarily enhance the signal-to-noise ratios for these phenotypes, as loss of power resulting from reporting error may equate to discarding up to half of the sample ^*^. Considerations on statistical power and sample size requirements should therefore not only focus on the genetic architecture of the trait and the study design, but also incorporate phenotype resolution as a parameter of interest.

Examining factors contributing to reporting error, we found that reporting error varied systematically across sociodemographic groups. In particular, young, female participants with higher intelligence scores and those from a socio-economic favourable background (higher education and income) tended to provide the most accurate self-report information. This is consistent with the notion of heteroskedastic error, where the error variance depends on certain sample characteristics (e.g., the accuracy in reporting level of education depends on education itself, cf. **Figure 5A**). The impact of this error structure on study findings will depend on the research question of interest; if gene-discovery is the main goal, error in the phenotype reduces power and increases Type-II error rates. While increasing the sample size (i.e., reduced sampling error) could compensate for the loss of power, such efforts would not correct for the downward bias in estimates of variance components (e.g., SNP heritability, polygenic prediction) resulting from error in the phenotype. For example, for phenotypes with high levels of reporting error, we observed relative *h*^*2*^-attenuation of up to 20% (cf. **Figure 6A**). As such, part of the missing heritability problem results from poor phenotype ascertainment, such as the use of minimal phenotyping or misclassification1. Similarly, the higher *h*^*2*^ observed for physical attributes (e.g., height, eye colour) than for socio-behavioural traits (e.g., smoking, SES) in the UKBB^23^ may not solely reflect a stronger genetic component, as measurement problems are mostly inherent to the latter traits.

In classical observational analyses, bias will occur if reporting error is present in the exposure, which attenuates effect estimates towards the null (cf., regression dilution or attenuation bias^24,25^). In this scenario, the bias on parameter estimates can be particularly large, potentially exceeding bias resulting from other sources (e.g., selective participation, **Figure 6B**). As such, while large-scale biobanks are imperative for the study of biological pathways of small effects, such minimally phenotyped convenience samples may not be a strong contender for classical (non-genetic) epidemiological research. For that, smaller but more representative samples with gold-standard measures are the potentially more trustful alternative.

Finally, we compared features underlying reporting error to those of other participation behaviours, here the UKBB participation propensity. We found that individuals with high self-report quality were more likely to participate in the UKBB, and that the application of statistical tools designed to ensure sample representativeness (probability weighting) increased self-report errors. This finding is consistent with findings from survey research, where probability (i.e., representative) samples showed more measurement error than volunteer samples^26^, and where efforts to enhance data quality reduced sample representativeness^27,28^. Together, these results highlight that biases resulting from response and participation behaviours are not independent and operate in opposite directions, such that adjusting for one type of bias could aggravate bias resulting from other sources.

Consequently, design considerations should also focus on finding an optimal trade-off between sampling bias and phenotype precision. For example, the application of reporting error (inverse-variance) weights enhanced phenotype resolution in the UKBB without further compromising the level of representativeness in the UKBB (**Figure 4**). Collecting quality indicators and metrics for phenotype precision (e.g., use of tools to screen for poor questionnaire responding^29^) in future biobanks may therefore prove useful to remove some of the noise in the phenotype.

A key consideration when interpreting our results relates to the error structure examined here. More specifically, our work focused on inconsistent self-reporting over time (i.e., random fluctuations in the phenotype), rather than sources of consistent misreporting (i.e., systematic over- or underreporting, cf. **sFigure 1D**, Supplement). Systematic error, documented for numerous traits (e.g., self-reported weight, where overweight individuals tend to underreport^30^), can only be explored if error-free reference data is available. For that reason, it was also not possible to explore error in phenotypes subject to temporal variability (e.g., self-reported alcohol use), as the data at hand did not allow us to distinguish reporting error from environmental influences on the observed within-individual variability. Finally, the reporting error mechanisms identified in this work may not translate to other cohorts, as differences in recruitment schemes and population characteristics likely impact how error in self-report measures is expressed.

In summary, our findings emphasize that both self-report data quality and sampling features are potential sources of poor reproducibility for biobank-scale research, leading to imprecision and bias that can complicate the interpretation of findings. Analogous to quality control procedures developed for the processing of genetic data, the application of tools designed to enhance phenotype resolution (e.g., repeat measurements, regression calibration^16^, imputation^31^, weighted regression) and sample representativeness (e.g., probability sampling or weighting) should therefore become an integral part of data collection, pre-analytic data handling and sensitivity checks.

## Data availability

The reporting error genome-wide association statistics will be made available through the GWAS catalog.

## Code availability

The following software was used to run the analyses:

REGENIE (https://github.com/rgcgithub/regenie)

TwoSampleMR (https://mrcieu.github.io/TwoSampleMR/)

GenomicSEM (https://github.com/GenomicSEM/GenomicSEM).

All analytical scripts are available at https://github.com/TabeaSchoeler/TS2023_repErrorUKBB.

## Acknowledgements

This research has been conducted with the UK Biobank Resource under application number 16389; we thank all biobank participants for sharing their data. This study would not have possible without the use of publicly available genome-wide summary data and software tools. The authors gratefully acknowledge these resources, and thank the research participants, the research teams and institutions that have contributed to this research. Computations have been performed on the HPC cluster of the Lausanne University Hospital.

## Footnotes

* Assuming an *r*^2^_TM_ of 0.5, where *r*^2^_TM_ is the square of the correlation between the true phenotype (*p*_T_) and the measured phenotype (*p*_M_)^32^ and n is the sample size.

