## Supplement file for "Self-report inaccuracy in the UK Biobank: Impact on inference and interplay with selective participation"

### sFigure 1. Impact of error in the phenotype on genotype-phenotype associations

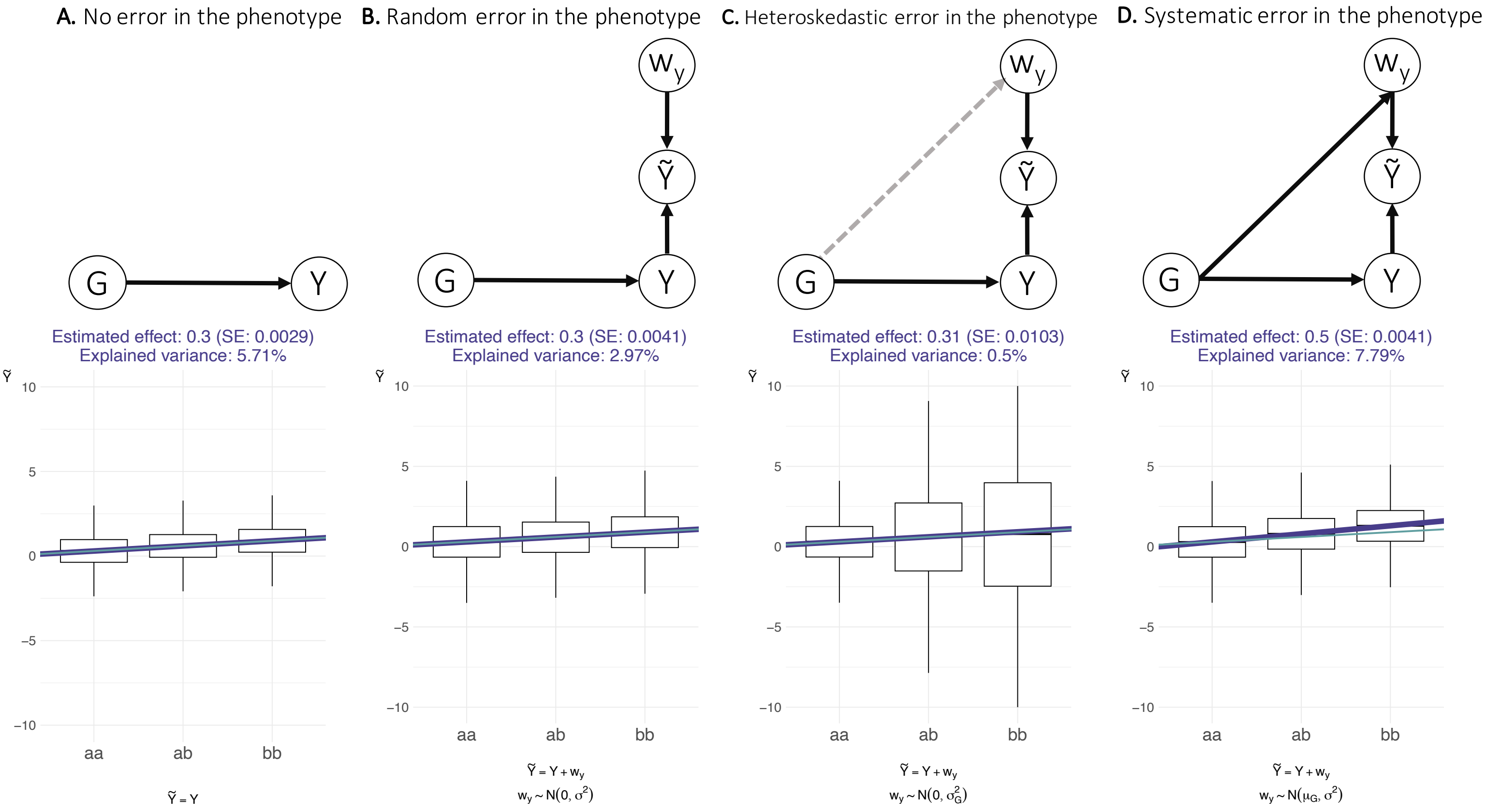

Illustration of possible measurement error mechanism when testing the effect of a single genetic variants (G, assumed to be measured without error) on a continuous phenotype measured without error (Y) or with error ($\tilde{Y}$). $\tilde{Y}$, the observed phenotype, is a function of the true value of the phenotype (Y) and the error in the phenotype ($w_y$), i.e., $\tilde{Y} = Y + w_y$. Biallelic SNP genotypes (aa, ab, bb) are presented on the X-axes. Random error in the phenotype (illustrated in **panel B**) occurs when $w_y$ is unrelated to G and Y, here simulated by adding a random normal variable with a mean of zero and a standard deviation of one (i.e., constant or homoskedastic error variance) to the model. Random error in the phenotype does not induce bias in SNP effects, but increases the standard errors and reduces the variance explained by the genotype. Heteroskedastic error in the phenotype (illustrated in **panel C**) occurs when the variance in $w_y$ depends on G (e.g., where bb-carriers show larger random errors than aa-carriers, i.e., error that is not constant across G). Heteroskedastic error does not induce bias in the SNP effects, but leads to incorrect standard errors. Systematic error (i.e., error that is not random and where the direction of the error is different across G, illustrated in **panel D**) occurs if the mean in $w_y$ depends on G ($\mu_{aa}$=0, $\mu_{ab}$=0.2, $\mu_{bb}$=0.4). Systematic error results in biased SNP estimates.

#### sFigure 2. Concordance between objective versus subjective measures

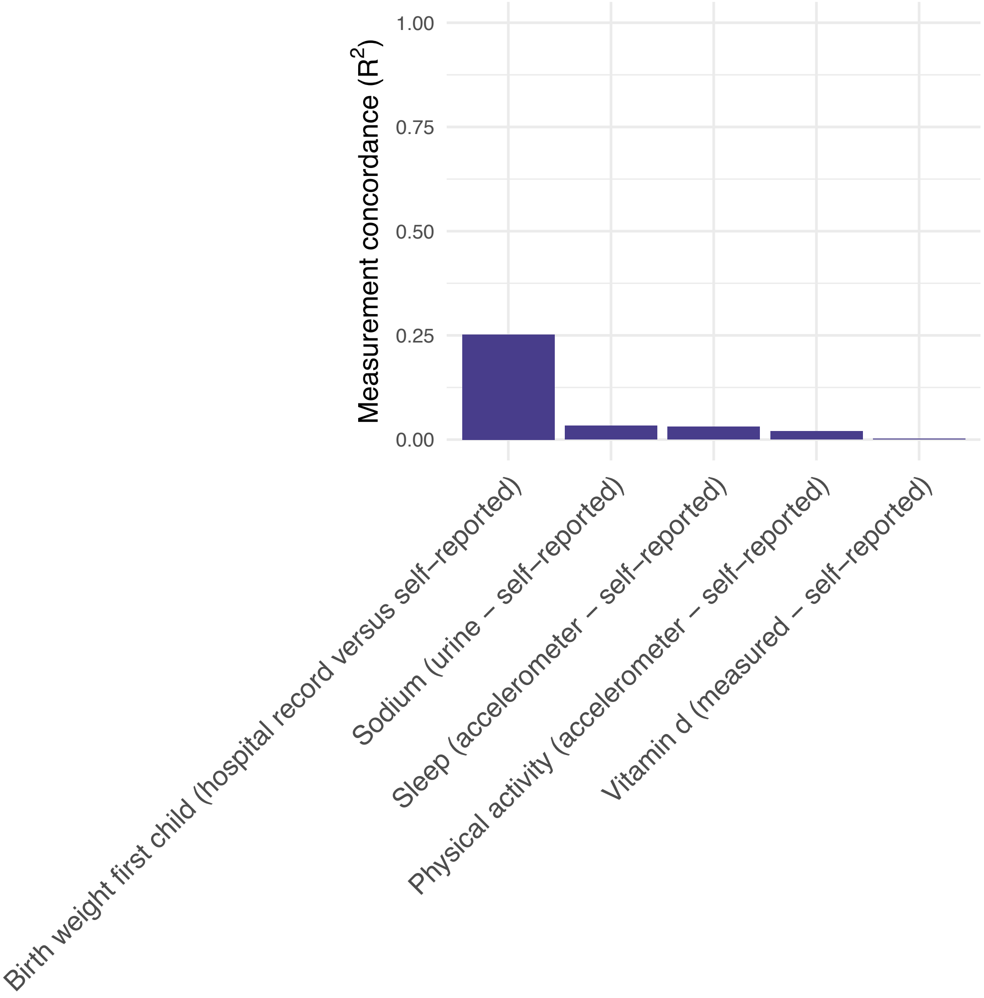

Measurement concordance ($R^2$) comparing objective and subjective measures for five UKBB phenotypes. $R^2$=Variance explained by models regressing objectively ascertained phenotype (e.g., accelerometer derived sleep duration) onto the subjectively ascertained phenotype (e.g., self-reported sleep duration), while controlling for follow up time. The following UKBB variables were used: Birth weight of the first child, including self-reported (ID 2744, 'what was the birth weight of your first child in pounds?') and hospital recorded (ID 41284, the birth weight of the first baby born to the participant in their hospital inpatient records); Physical activity (as done in previous studies[1]), including self-reported (ID 22040, the total Metabolic Equivalent Task minutes per week) and accelerometer derived (ID 90019-90025, used to derive an acceleration average. Individuals with poor wear time (ID 90015) were excluded); sleep duration, including self-reported (ID 1160, 'about how many hours sleep do you get in every 24 hours?') and accelerometer derived (ID 90027-90050, used to derive the average number of hours of sleep per 24 hours, applying a cut-off of $\leq$10 milligravity to indicate sleep. Individuals with poor wear time (ID 90015) were excluded); sodium intake, including self-reported (ID 26052, sodium intake obtained from the 24-hour dietary recall questionnaire) and measured (ID 30530, sodium measured in urine); vitamin D intake, including self-reported (ID 100021, vitamin D obtained from the 24-hour dietary recall questionnaire) and measured (ID 30890, vitamin D obtained from blood measures).

### sFigure 3. Effects of age and follow up duration on residual scores ($RES_i$)

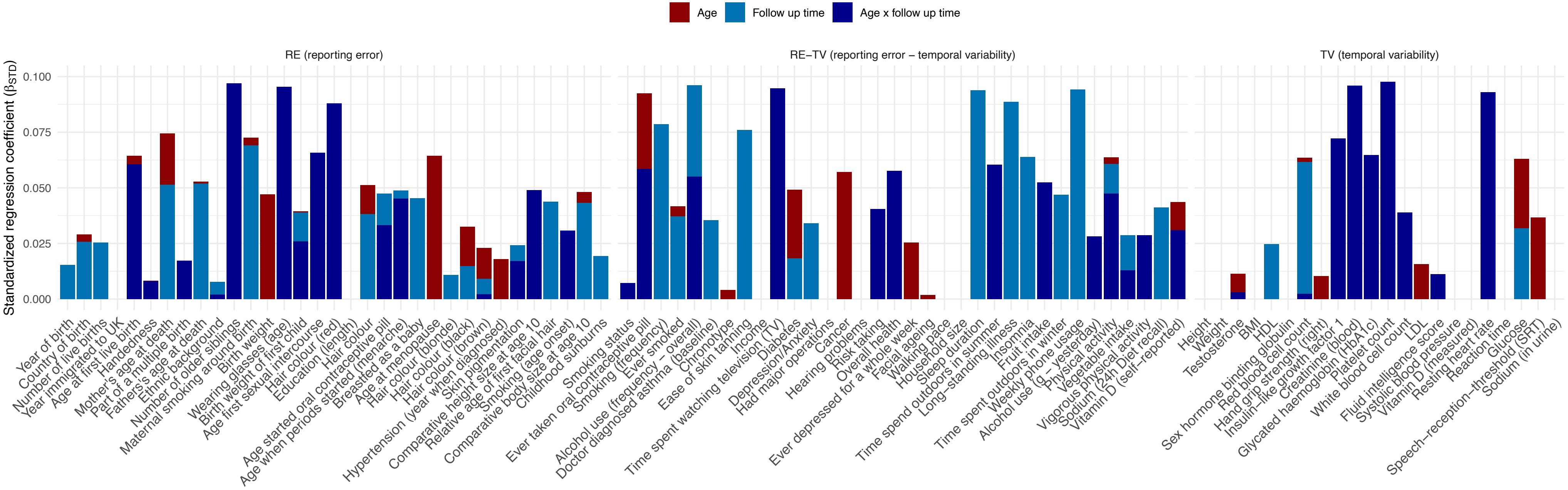

Standardized regression coefficients of age and follow up duration on the raw residual scores. $RES_i$ are derived from a model regressing the phenotype measured at time point 2 ($P_{T2}$, e.g., birth weight reported at follow up) onto the phenotype assessed at time point 1 ($P_{T1}$, e.g., self-reported birth weight assessed at baseline).

### sFigure 4. Sex differences in residual scores

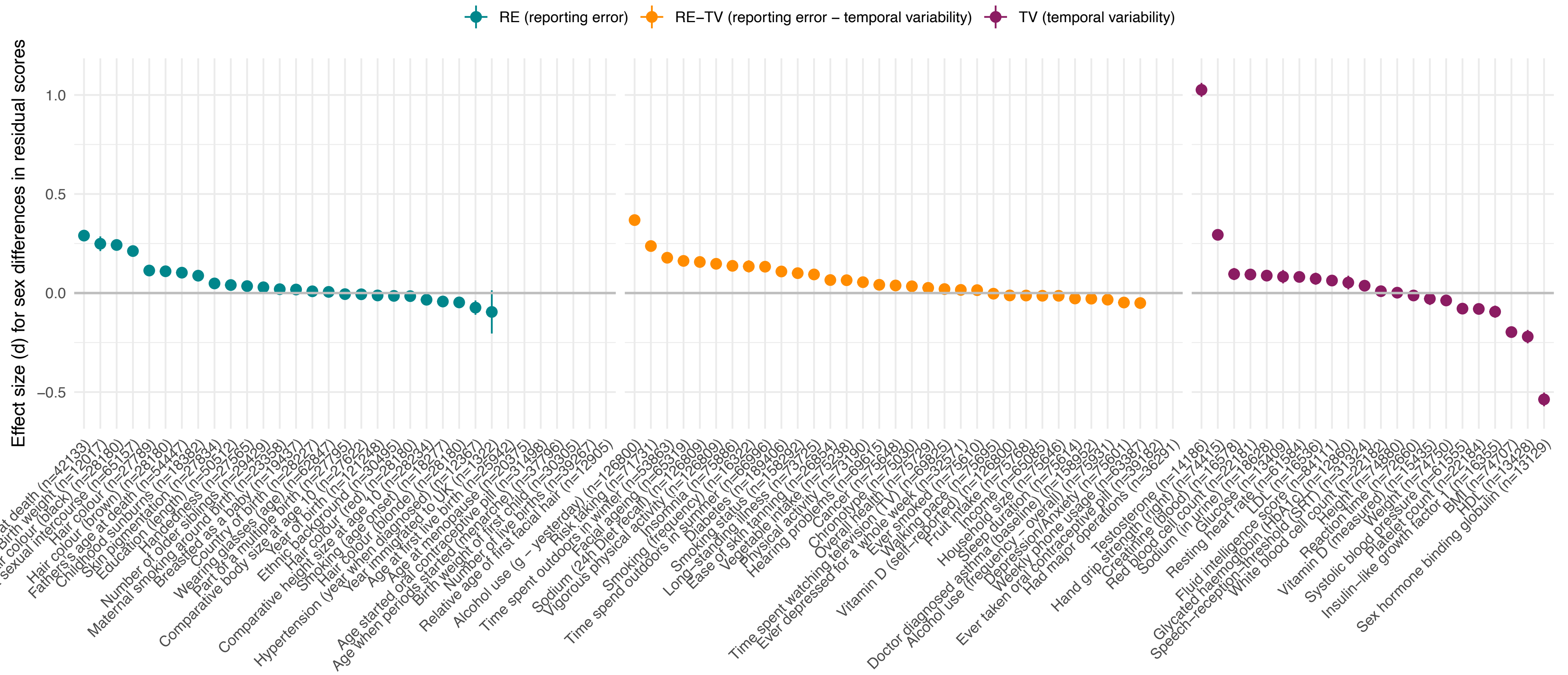

Estimates of Cohen's *d* quantifying sex differences in residual scores. The residual scores are derived from a model regressing the phenotype measured at time point 2 ($P_{T2}$, e.g., birth weight reported at follow up) onto the phenotype assessed at time point 1 ($P_{T1}$, e.g., self-reported birth weight assessed at baseline), while controlling for follow up time ($time_{T2-T1}$). Traits with missing estimates of Cohen's *d* index those that are assessed in either males (e.g., age of first facial hair) or females (e.g., age at menopause) only.

### sFigure 5. Phenotypic variance explained by genetic variants reaching genome-wide significance

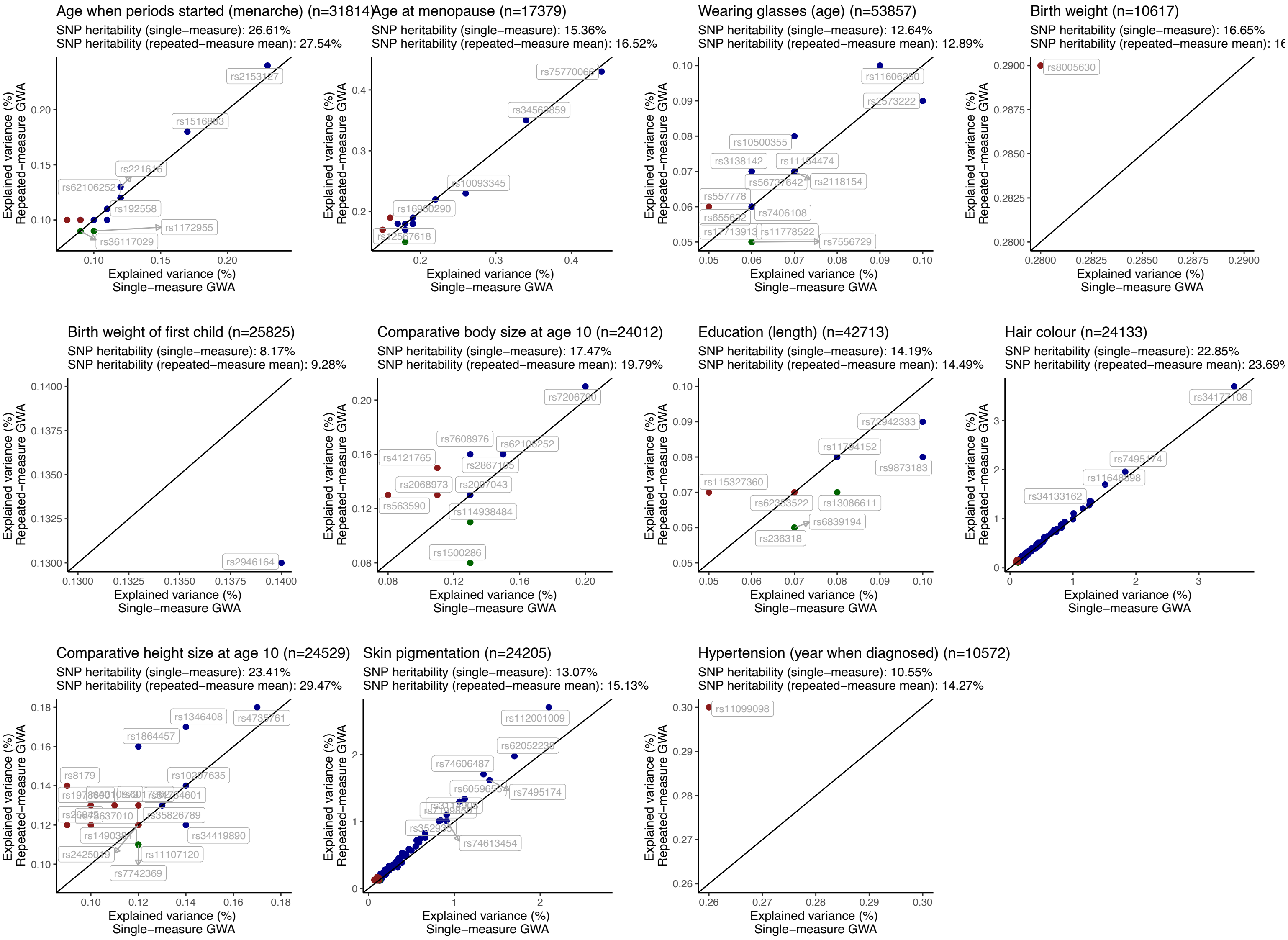

Explained variance ($\beta_{STD}^2$) per SNP reaching genome-wide significance in error-corrected GWA analyses (y-axis, phenotype obtained using means across multiple measurement occasions) or error-uncorrected GWA analyses (x-axis, phenotype obtained from a single baseline measure). The colour scheme highlights in which GWA the genetic variant was identified, including error-corrected GWA (in red), error-uncorrected GWA (in green) or in both (blue).
